# Bridging Data Gaps in Oncology: Large Language Models and Collaborative Filtering for Cancer Treatment Recommendations

**DOI:** 10.1101/2025.04.07.25325243

**Authors:** Tengjie Tang, Angkai Li, Xingye Tan, Qingli Ji, Lu Si, Le Bao

## Abstract

**Background:** Patients with rare cancers face substantial challenges due to limited evidence-based treatment options, resulting from sparse clinical trials. Advances in large language models (LLMs) and recommendation algorithms offer new opportunities to utilize all clinical trial information to improve clinical decisions.

**Methods:** We used LLM to systematically extract and standardize more than 100,000 cancer trials from ClinicalTrials.gov. Each trial was annotated using a customized scoring system reflecting cancer-treatment interactions based on clinical outcomes and trial attributes. Using this structured data set, we implemented three state-of-the-art collaborative filtering algorithms to recommend potentially effective treatments across different cancer types.

**Results:** The LLM-driven data extraction process successfully generated a comprehensive and rigorously curated database from fragmented clinical trial information, covering 78 cancer types and 5,315 distinct interventions. Recommendation models demonstrated high predictive accuracy (cross-validated RMSE: 0.49–0.62) and identified clinically meaningful new treatments for melanoma, independently validated by oncology experts.

**Conclusions:** Our study establishes a proof of concept demonstrating that the combination of LLMs with sophisticated recommendation algorithms can systematically identify novel and clinically plausible cancer treatments. This integrated approach may accelerate the identification of effective therapies for rare cancers, ultimately improving patient outcomes by generating evidence-based treatment recommendations where traditional data sources remain limited.

## 1 Introduction

Rare cancers, defined as those with fewer than six new cases per 100,000 people annually, collectively account for approximately 25% of all cancer diagnoses. Despite their substantial prevalence, rare cancers face unique challenges: a five-year overall survival rate of only 49%, significantly lower than the 63% observed for more common cancers. These disparities are attributed to small sample sizes, limited research resources, difficulties in early diagnosis, and fewer evidence-based treatment options. To overcome these obstacles, interdisciplinary collaboration and innovative data approaches are essential.

Recent advances in clinical research have generated an enormous volume of data from clinical trials, providing a valuable resource for improving cancer treatment. However, the unstructured and fragmented nature of clinical trial data – dispersed across numerous scientific articles and databases – creates significant barriers to its effective use. Extracting, processing and synthesizing this information into actionable insights remains a critical unmet need, particularly for rare cancers. Large Language Models (LLMs), with their exceptional ability to process natural language, clean and standardize unstructured data, and recognize complex patterns, offer a promising solution to these challenges. An LLM is a type of generative artificial intelligence (AI) model that is designed for tasks related to natural languages. OpenAI’s GPT, Meta AI’s LLama, Anthropic’s Claude, Mistral AI’s Mistral and Mixtral are examples of the current popular LLMs. LLMs can be “prompted” by a given task to perform natural language processing (NLP) tasks and generate human-like texts [1]. LLMs have significantly impacted various aspects of clinical trials and medical documentation [2, 3], including clinical trial planning [4, 5], patient-trial matching [6], technical writing [7], clinical decision making [8, 9], and medical queries [10, 11].

In this study, we propose a novel framework that integrates large language models (LLMs) with collaborative filtering techniques to enhance treatment decisions in oncology, particularly for rare cancers. Leveraging natural language processing capabilities, our approach systematically extracts and standardizes critical information from over 100,000 cancer-related clinical trials, resulting in a comprehensive and meticulously curated database. This structured dataset enables the development of advanced recommendation systems, including k-nearest neighbors, matrix factorization, and neural collaborative filtering, to identify promising treatments based on clinical trial outcomes. The recommendations generated by these models are further validated by cross-validations and oncology experts to ensure reliability and clinical relevance. By combining cutting-edge artificial intelligence methods with oncology knowledge, our framework effectively bridges the gap between clinical research findings and practical decision making in patient care. Ultimately, our approach not only seeks to improve clinical outcomes for patients with rare cancers, but also establishes a foundation for the broader integration of AI-driven methodologies into clinical research.

## 2 Method

### 2.1 Data Collection and Standardization

We collected data from ClinicalTrials.gov, a comprehensive database of clinical studies conducted world-wide. Up to 2024, it listed more than 100,000 cancer research studies; Each reports a trial-level summary, including the number of National Clinical Trials (NCT), study title, study status, type of cancer, study design, study type, interventions, phase of the trial, eligibility criteria, outcome, and the relevant date. In addition, we retrieved the study arms and the corresponding PMC IDs for the clinical trials using APIs provided by ClinicalTrials.gov.

Since the summaries were manually entered by the research teams conducting the trials, drug names and cancer types often appear in non-standardized formats, and errors may have been introduced during data entry. Furthermore, most trials do not include information on the effectiveness of treatment, limiting the scope of available data. As high-quality, standardized information is critical for advanced statistical analyses, additional data cleaning and processing steps were required. We used Llama 3 in the HuggingChat API to extract key information from extensive clinical trial studies presented in textual format, as detailed below.

#### 2.1.1 Prompt to obtain the results of clinical trials

We obtained abstracts of the publications that contained PMC IDs and excluded meta-analyses that did not report specific clinical trial results. We then applied the LLM to analyze these filtered abstracts. The model evaluated the conclusions for each experimental arm to determine whether the treatment showed promise. The following is the prompt submitted to HuggingChat to evaluate whether each arm yielded promising results. The underlined sections indicate the information provided by the user.

“Please summarize the result of this text: <u>abstract</u> in the following format:

1. Specific conclusions for each of the following <u>number of arms</u> arm(s): <u>arm information</u>. Use single-level bullet point ‘•’ for each arm in order. You do not need to consider dose information. If you could not find, answer not applicable.
2. Whether the treatment is promising or not. Use single-level bullet point ‘•’ for each arm in order with one single keyword among ‘promising, not promising, unknown’ to summarize the conclusions you found in the subtitle 1. You do not need to consider dose information. If you cannot find results, say no results. There is no need to provide extra justifications.”

Here is an example of the text generated from LLM using the prompt:

- The combination of YM155 with alemtuzumab demonstrated therapeutic efficacy and markedly additive antitumor activity.
- Promising

Figure 1 (a) illustrates the data processing procedures used to determine the promising status of each study and its corresponding arms. To facilitate statistical analysis, we performed additional data processing to convert text-based results into a structured tabular format. In particular, we first converted the text file into a CSV format and then standardized the names of the treatments and cancer types.

**Figure 1:**
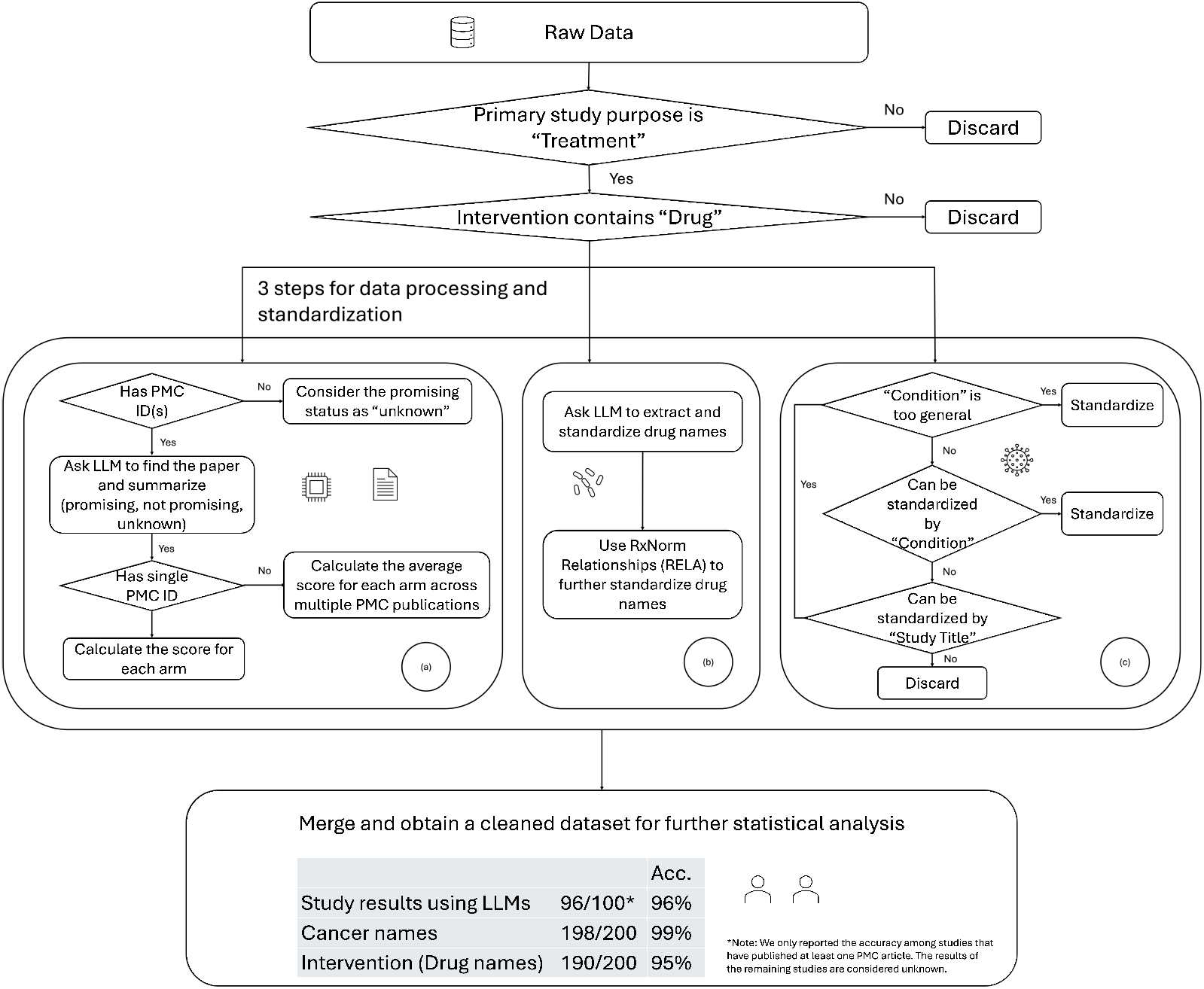
Data collection and processing pipelines: (a) obtain the study results based on the abstract of PMC articles, (b) standardize drug names, (c) standardize cancer types.

#### 2.1.2 Prompt to standardize the drug names and cancer names

The intervention column in our CSV dataset contains drug information for each clinical trial arm. These entries often include varying formats and details such as the use of brand names versus generic names, the use of abbreviated forms, combination treatments, different formatting styles, variation in dosage representation (e.g., “Omaveloxolone Capsules (2.5 mg/capsule)” or “Nivolumab (240 mg)”), and descriptive treatment descriptions (e.g., “3-year abemaciclib without chemo”). To address these challenges, we first developed the following prompt to extract drug names from raw data:

> “Identify all drugs in this entry: <u>the intervention information of a particular clinical trial arm</u>, and only output the drug names separated by comma. If you cannot find a drug, ouput NA. Please do not include additional information like doses or explanation.”

We also employed natural language processing (NLP) techniques to validate the precision of the drug name. To capture variations and slight discrepancies between treatments in raw data and those extracted via LLM, we implemented several matching methods, including fuzzy matching, TF-IDF cosine similarity, and Word2Vec semantic similarity [12]. When the similarity between two drug names exceeds a predefined threshold, they are considered equivalent, achieving an accuracy of 97% across all clinical trials. Finally, we reran our drug information prompt for those treatments with a mismatch determined by NLP. This additional round of information extraction further improves the reliability of our standardization approach. We further standardize drug names using the RxNorm dataset [13], developed by the National Library of Medicine (NLM), which documents relationships among different drug terminologies. Specifically, we determine whether two drugs are synonyms or whether one is a brand name and the other is a generic name, or whether one drug is a special form of another drug based on RxNormal (RELA) relationships. Figure 1 (b) outlines the pipeline used to standardize drug names.

Finally, Figure 1 (c) illustrates our approach to standardizing cancer names. The Initial dataset entries occasionally contained overly broad descriptors, such as “advanced solid tumor” or “adult cancer.” These general terms were excluded from further analysis, as retaining them could decrease recommendation precision. Using a comprehensive cancer classification list from the National Cancer Institute (NCI), we used LLMs to identify synonyms and subtypes, subsequently compiling these into a standardized dictionary of 99 distinct cancers or tumors. Appendix Table A1 summarizes the 25 most frequent and least frequent cancers captured in our dataset. To enhance completeness, we also searched study titles for possible cancer types that were absent from the “Condition” entry in the ClinicalTrials.gov database. Entries lacking dictionary matches were removed. Most of the unstandardized conditions excluded at this stage were non-cancerous conditions (see Appendix Table A2).

#### 2.1.3 Dataset for Recommendation Purposes

For our original data set, we have 23,224 Phase I trials, 37,152 Phase II trials, 10,309 Phase III trials and 2,457 Phase IV trials. Of these 73,142 trials, 9,719 of them are in multiple phases and we only kept the trials in the highest phase. For treatment recommendation purposes, we excluded trials with missing cancer or intervention information, trials whose primary study purpose was not treatment or did not involve any drug. We also discarded 3,196 trials with overly general conditions and 5,007 trials lacking standardization, primarily because they were not related to cancer or tumors (see Appendix Table A2). The final dataset contains 20,769 trials.

During the modeling process, trials involving multiple cancer types were separated into individual [cancer, treatment] pairs. Cancer types associated with fewer than 20 unique treatments were excluded, following common practice in collaborative filtering [14]. This resulted in a final dataset of 35,425 pairs, comprising 78 distinct cancer types and 5,315 distinct drugs. To ensure accuracy, we manually reviewed 100 randomly selected trials with at least one published PMC article, and 100 randomly selected trials without publications from this final dataset. We found a 96% accuracy in study results among trials with publications, and observed correctness rates of 99% for cancer names and 95% for intervention names across 200 test cases, respectively.

### 2.2 Scoring System

Leveraging a comprehensive and structured cancer clinical trial database, we created a scoring system to evaluate the interactions between cancers and treatments. This scoring system incorporates factors such as clinical outcomes, trial stages, experimental or control status of the trial arm, and trial year.

- We use the trial phase number (i.e., Phase I, II, III, or IV) as a baseline score. Higher-phase trials are typically more promising than lower-phase trials.
  - If the trial status is “terminated”, “suspended,” or “withdrawn”, we reduce the baseline score by one. It steers recommendations away from treatment paths that are likely associated with safety concerns, lack of efficacy, or logistical issues.
  - For the “completed” trials, if the results are not promising, we also reduce the baseline score by one; if the arm has promising outcomes or is a control arm, we keep the baseline score as the trial phase number. This ensures that clinically vetted (promising) or standard-of-care (control) options are viewed more favorably than experimental trials that do not yield promising results.
  - When the outcome is unknown, the score is set to “phase *−*0.5” as a moderate step between promising and unpromising.
- Medical treatment standards evolve rapidly, and newer clinical trials often reflect the latest advances. To account for this, we subtract 0.05 *×* (2020-year) from the score if the clinical trial was conducted before 2020, giving less weight to older trials. This adjustment ensures that the recommended treatments remain relevant and in line with current medical standards.

After data cleaning, we obtained a tabular dataset that includes each cancer type, its associated treatment, the clinical trial year, and the corresponding score. Each data point represents a single cancer type and a single trial arm; if a trial arm is linked to multiple cancer types, it is divided into separate data points. The scores range from −1.7 to 4 across all cancer-treatment combinations, with first-quartile, median, and third-quartile values of 0.5, 1.15, and 1.5, respectively.

### 2.3 Recommendation System

A recommendation system is a computational framework designed to predict user preferences and suggest relevant items—such as products, movies, or articles—by analyzing data patterns. In this study, we developed a recommendation system specifically designed to identify promising treatments for various cancer types.

Assume that we have a set of users of users *u* = 1, *· · ·, U* and items *i* = 1, *· · ·, I*. Let *y*_*ui*_ denote the score made by the user *u* on the item *i*, and *ŷ*_*ui*_ represent the predicted score, which can be roughly interpreted as the clinical trial stage at which a new cancer treatment combination is likely to reach. In Collaborative Filtering (CF), the term **Collaborative** refers to the use of similarities between users or items to make predictions. The term **Filtering** refers to the process of recommending relevant items while excluding those that are not. CF approaches provide user-specific recommendations of items based on scores without other information about items or users [15]. The user-based CF recommends items based on the preferences of similar users. For example, if Brian and Bryson are identified as similar users and Brian has watched a particular show, Bryson may receive a recommendation for that show. On the other hand, the item-based CF suggests items similar to those in which a user has already shown interest. For example, if a person has watched *Family Guy*, the system might recommend *The Simpsons* due to their similarities.

We introduce three main collaborative filtering (CF) approaches: *k*-nearest neighbors, matrix factorization, and neural collaborative filtering. The objective is to predict treatment scores for cancers that currently lack corresponding clinical trials.

#### 2.3.1 *k*-Nearest Neighbor

The neighborhood-based approach focuses on relationships either between items or between users. For example, it can model a user’s preference for an item based on the user’s ratings for similar items. A *k*-NN classifier identifies the *k* closest points from the training data and assigns class labels based on these nearest neighbors.

In the user-based neighborhood approach, the predicted score *ŷ*_*ui*_ for user *u* on a new item *i* is derived from the scores assigned by the *k* users most similar to *u* on the item *i*. Let 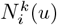 represent the set of up to *k* neighbors with the highest similarity to the user *u*. Then, the prediction *ŷ*_*ui*_ is calculated as:

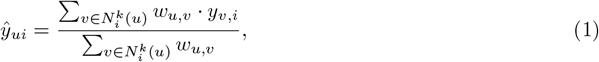

where *w*_*u,v*_ is a similarity measure between user *u* and *v*. We use the cosine similarity in the work.

Equation (1) is called the user-based neighborhood method. There is also an item-based method that recommends new and similar items to users based on the user’s preferences for existing items. However, it has been shown that if the number of users is less than the number of items, the user-based neighborhood method is superior [15].

#### 2.3.2 Matrix Factorization (MF)

The latent factor approach transforms users and items into a shared latent factor space and has gained popularity following its success in the Netflix Prize Competition [16, 17]. The recommendation problem is formulated as a matrix completion task: users correspond to matrix rows, items to columns, and entries represent user-item interactions. However, real-world data are typically sparse — most users only interact with a small subset of available items. In the Netflix Prize dataset, for example, approximately 99% of the ratings are missing because users rate only a small fraction of the available movies. The latent factor approach addresses this sparsity by approximating the original user-item matrix with a set of lower-dimensional latent factors. This abstraction allows the model to generalize from limited data, capturing underlying patterns that explain user preferences and item characteristics.

Matrix factorization (MF) is a popular latent factor technique that maps both users and items to a joint latent space of dimension *K*. Let **p**_*u*_ ∈ ℝ^*K*^ represent the latent vector for user *u* and **q**_*i*_ ∈ ℝ^*K*^ represent the latent vector for item *i*. MF then estimates the interaction between user *u* and item *i* as the inner product:

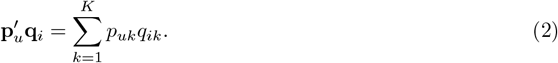

We predict the rating by

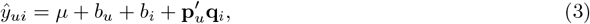

To obtain estimates of the unknown parameters, we minimize the regularized sum of squared errors:

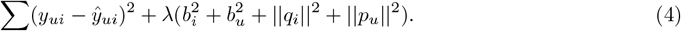

Empirical studies show that MF often outperforms simpler models by uncovering hidden user-item patterns [14]. However, as a linear model that relies on inner products, it may fail to capture more complex user interaction structures.

#### 2.3.3 Neural Collaborative Filtering (NCF)

He et al. 2017 [14] introduced Neural Collaborative Filtering (NCF) to replace the simple inner product of MF with a flexible deep neural architecture. This allows NCF to model complex, nonlinear interactions and outperform traditional MF approaches. It can also seamlessly integrate additional information (e.g., gene mutations) for more accurate recommendations.

Within our Neural Collaborative Filtering (NCF) framework, the process begins by transforming feature vectors representing cancers and interventions into dense embeddings that capture their underlying relationships. These learned embeddings serve as inputs to neural collaborative filtering layers that predict treatment scores by minimizing discrepancies between predicted and actual outcomes. We use the Neural Matrix Factorization (NeuMF) model to predict scores for all [cancer, treatment] pairs. It combines two components: one that captures interactions through a simple matrix factorization method, and the other that uses a multilayer perceptron to learn more complex patterns. The embedding dimensions are carefully chosen to balance model complexity and performance. Finally, we use the following strategies to improve the robustness of the model and prevent overfitting: (1) *l*_2_ regularization to mitigate noise in training data, (2) optimization of the adaptive learning rate to improve convergence efficiency, and (3) early stopping criteria to stop training when validation performance plateaued.

## 3 Results

We perform hyperparameter tuning with extensive grid searches for *k*-NN and MF (with SVD implementation), while employing a random search with a predetermined number of trials for the NCF model. The root mean squared errors (RMSE) are 0.622, 0.605 and 0.491 for *k*-NN, MF, and NCF, respectively. The predicted scores indicate the plausible phase number that a new treatment could reach, and the RMSE results show that the prediction will likely be off by half of a phase number.

Evaluating recommendations for rare cancers is challenging with limited empirical evidence; therefore, we use the melanoma treatment recommendation as an illustrative example. Melanoma represents only a small fraction of cases of skin cancer but causes most skin cancer-related deaths due to its aggressive nature. In 2024, among the clinical studies developed for other cancer types, our top recommendation for melanoma patients is to adopt the OPTI-DOSE clinical trial (NCT05949424), which investigated dosing strategies for oral anticancer drugs in older carcinoma patients. It started treatment at lower doses than the drug label with escalation based on tolerability to enhance treatment tolerability while maintaining efficacy. All five drugs evaluated in the study have been documented to be used in the treatment of melanoma. Olaparib is a PARP inhibitor, and emerging evidence suggests that combining Olaparib with BRAF and MEK inhibitors may benefit melanoma patients with BRAF mutations [18]. Palbociclib, a CDK4/6 inhibitor that blocks cell cycle progression, has also shown promise in melanoma treatment [19, 20]. Furthermore, tyrosine kinase inhibitors such as lenvatinib, sunitinib, and pazopanib, which target various pathways, have been explored for treating melanoma patients [21, 22].

Finally, we provide the data and code designed for quick and flexible implementation of recommendations. By downloading the processed dataset, trained models, and the best hyperparameters, users can instantly generate recommendations. The code allows for customization of the input cancer type and the number of top interventions to recommend. This flexibility ensures that users can easily adjust the cancer type for which they want recommendations and specify how many top-ranked interventions to retrieve, enabling rapid, tailored outputs without re-training the model.

## 4 Discussion

In this study, we demonstrated that large language models can be harnessed to streamline the extraction and utilization of clinical trial data. Starting with more than 100,000 cancer trials listed on ClinicalTrials.gov, we used an LLM to extract outcome data from thousands of trial publications and created a structured dataset of nearly 21,000 trials (encompassing 78 cancer types and 5,315 drugs) annotated with their results. Using this resource, we developed a collaborative filtering framework that recommends potentially effective treatments across cancer types. The model identified several treatment regimens originally tested in other cancers as top candidates for melanoma, illustrating the utility of new treatment option suggestions for hard-to-treat cancers.

This work offers a novel data-driven approach to address persistent challenges in rare cancer research. A major strength of our approach is the integration of an LLM for data curation: the model achieved high precision in extracting trial arm results and standardizing terminology, overcoming the heterogeneity in how trial outcomes are reported. This allowed us to aggregate a far larger evidence base than any individual clinician or researcher could manually synthesize. Furthermore, by incorporating the trial phase, outcomes, and other context into a scoring system, we ensured that the recommendation model considers clinical relevance (for example, prioritizing successful Phase III and IV interventions over early phase or failed approaches).

To our knowledge, this study is the first to generate treatment recommendations using a comprehensive clinical trial dataset systematically curated by LLM. The identification of an immunotherapy–angiogenesis combination (camrelizumab plus thalidomide with chemotherapy) from a lung cancer trial for potential use in melanoma aligns with the emerging understanding of tumor immunology, yet was discovered automatically by the model that scans thousands of trials outside of melanoma. If refined and validated, such a system could serve as an ever-learning knowledge base to guide oncologists, especially when faced with a rare cancer patient who has limited treatment options.

Improving the interpretability of the model’s output is a natural next step to allow clinicians to understand why a particular treatment is recommended. Lastly, as new trial results and more powerful LLMs and recommendation systems become available, our framework will need to be continually updated.

## Data Availability

All data produced in the present work are contained in the manuscript.

## 5 Acknowledgment

Le Bao and Tengjie Tang were supported by NIH/NIAID R01AI136664 and R01AI170249. Lu Si was supported by National Natural Science Foundation of China (82425047,82272676); Beijing Natural Science Foundation (7242021); Beijing Municipal Administration of Hospitals’ Ascent Plan (DFL20220901). We extend our gratitude to Yuxuan Wu for her assistance in collecting and standardizing clinical trial data. This product uses publicly available data courtesy of the U.S. National Library of Medicine (NLM), National Institutes of Health, Department of Health and Human Services; NLM is not responsible for the product and does not endorse or recommend this or any other product.

## A Appendix

**Table A1:**
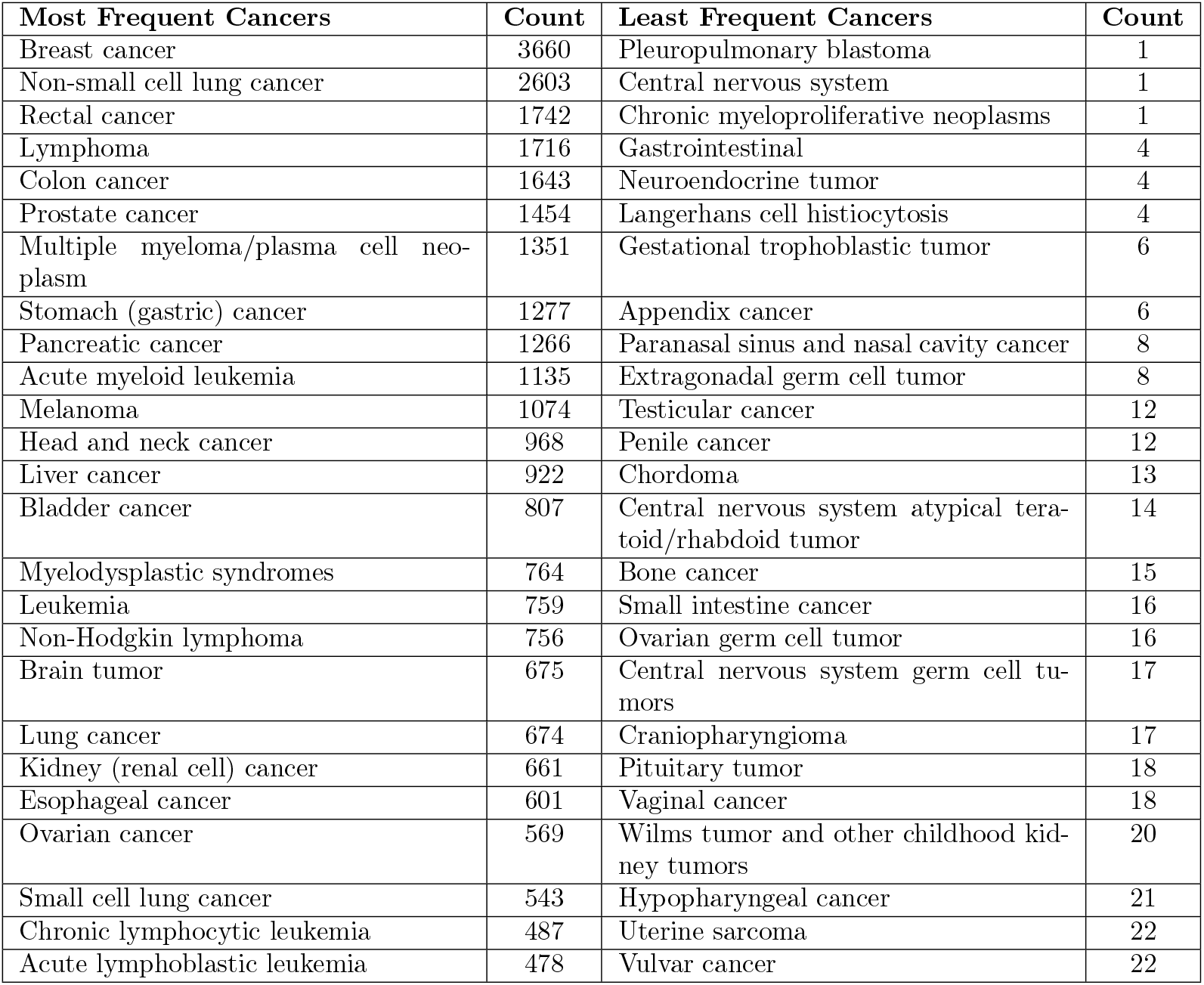
Top 25 Most Frequent and Least Frequent Cancers/Tumors (Cancers/Tumors with frequency less than 20 are removed from the final dataset.)

**Table A2:**
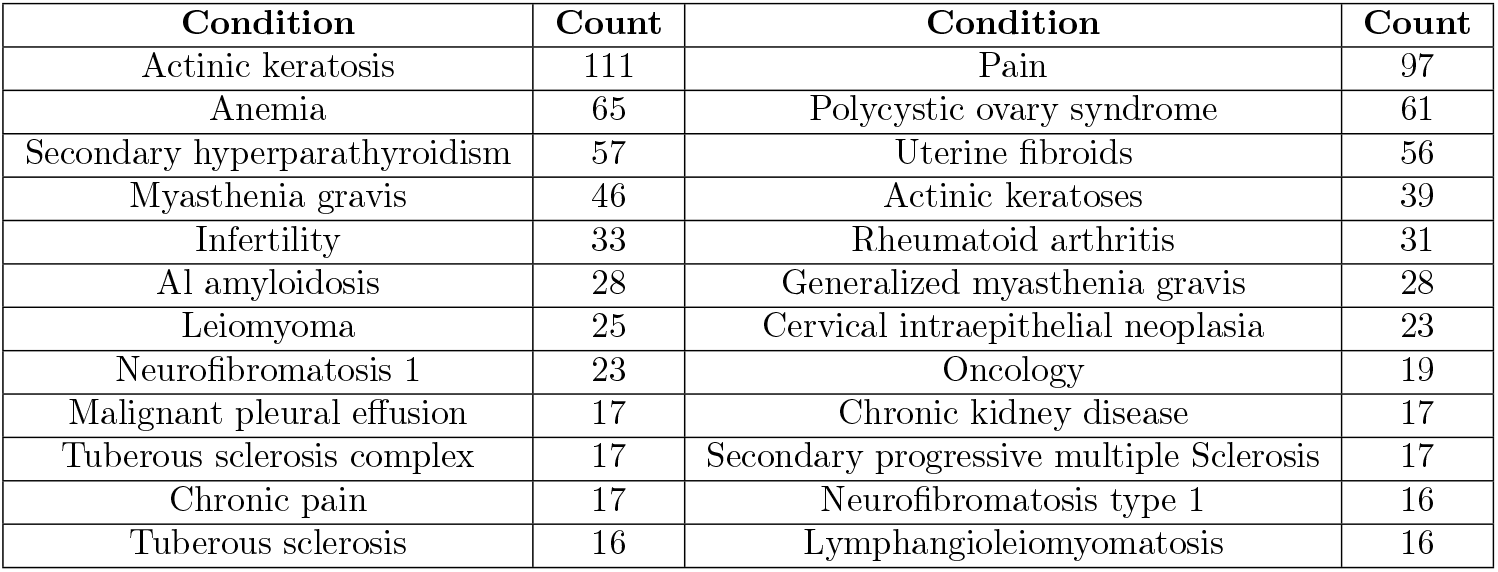
Conditions that are not successfully classified by our pipeline.

